# Robust immune response to the BNT162b mRNA vaccine in an elderly population vaccinated 15 months after recovery from COVID-19

**DOI:** 10.1101/2021.09.08.21263284

**Authors:** Hye Kyung Lee, Ludwig Knabl, Ludwig Knabl, Sebastian Kapferer, Birgit Pateter, Mary Walter, Priscilla A. Furth, Lothar Hennighausen

**Affiliations:** National Institute of Diabetes, Digestive and Kidney Diseases, Bethesda, MD 20892, USA; TyrolPath, Zams, Austria; Krankenhaus St. Vinzenz, Zams, Austria; Division of Internal Medicine, Krankenhaus Kufstein, Kufstein, Austria; Dr. Pateter’s surgery, Fliess, Austria; Clinical Core, National Institute of Diabetes, Digestive and Kidney Diseases, Bethesda, MD 20892, USA; Departments of Oncology & Medicine, Georgetown University, Washington, DC, USA

**Author notes:** Equal contribution (Hye Kyung Lee, Ludwig Knabl). Corresponding authors: HKL; LK; PAF; LH.

## Abstract

Knowledge about the impact of prior SARS-CoV-2 infection of the elderly on mRNA vaccination response is needed to appropriately address the need for booster vaccination in this vulnerable population. To address this, we investigated antibody and genomic immune responses in 16 elderly (avg. 81 yrs.) individuals that had received a single booster dose of BNT162b vaccine 15 months after recovering from COVID-19. Spike-specific IgG antibody levels increased in each of the study participants from an average of 710 U/ml prior to the vaccination to more than 40,000 U/ml within ten weeks after the vaccination. In contrast, anti-spike-specific IgG antibody levels averaged 2,190 U/ml in 14 healthy SARS-CoV-2-naïve individuals (avg. 58 yrs.) ten weeks after the second dose of BNT162b. RNA-seq conducted on PBMCs demonstrated the activation of interferon-activated genetic programs in both cohorts within one day. Unlike their transient induction in the younger naïve population, persistent activity and the initiation of additional cell cycle regulated programs were obtained in the older COVID-19 recovered population. Here we show that the elderly, a high-risk population, can mount a strong antibody and a persistent molecular immune response upon receiving a single dose of mRNA vaccine 15 months after recovery from COVID-19.

## Introduction

BNT162b mRNA COVID-19 vaccines have been shown to elicit strong antibody responses^1,2^ and be highly efficacious in real world settings^3^. While current research focuses on the long-term effectiveness and waning immunity^4^ in real world settings^5^, only few studies on vaccine efficacy in the elderly population have been reported^6,7^. Immunosenescence, defined as age-related decline of the immune system, has been associated with poor vaccine responses in older adults^8^ and previous studies have reported a negative relationship between age and mRNA vaccine-induced antibody titers^9,10^. However, recent data suggest that two doses of BNT162b elicit high IgG levels and serum neutralization in a SARS-CoV-2 antigen-naïve elderly population^7^. While optimal increases in antibody levels in SARS-CoV-2 antigen-naïve individuals requires two mRNA vaccine doses, individuals with prior SARS-CoV-2 infection have naturally acquired immunity and a single dose of vaccine is likely sufficient to boost antibody responses^10-12^. Yet, these studies have been conducted on younger cohorts, mainly health care workers, and the immune response of elderly COVID-19 recovered individuals (> 70 years) to a single dose of mRNA vaccines remains to be understood.

Studies on SARS-CoV-2 antigen-naïve and COVID-19 recovered subjects inoculated with mRNA vaccines have largely focused on measuring binding and/or neutralizing antibodies as primary end points. However, there is a definitive need to also understand the molecular immune responses after vaccination, both in COVID-19 recovered and SARS-CoV-2 antigen-naïve individuals.

Here we have addressed these knowledge gaps and assessed the immune response in an elderly population in a convent that experienced a SARS-CoV-2 outbreak in the spring of 2020. Sixteen nuns (avg. 82 yr.) that had recovered from COVID-19, received the primary booster vaccination 15 months after the initial infection. We assessed their antibody response and genomic responses through RNA-seq of peripheral blood mononuclear cells (PBMCs). As control, we investigated 14 SARS-CoV-2 naïve individuals (avg. 58 yr.) receiving two doses of BNT162b.

This real-world study provided critical information on the shared and unique molecular vaccine response and antibody production in two distinct groups, elderly individuals that were vaccinated 15 months after recovering from COVID-19 and naïve individuals receiving the two-dose regimen.

## Results

Here we investigated the immune response to the BNT162b vaccine of an elderly population recovered from COVID-19. A convent experienced a COVID-19 outbreak in the spring of 2020 and 16 nuns (average age 81.3 yrs.) with age-related underlying health conditions participated in this study (Fig. 1a; Supplementary Table 1). These individuals received a single dose of BNT162b dose approximately 15 months after testing positive for SARS-CoV-2 and developing mild cases of COVID-19. In parallel we investigated the immune response to BNT162b in a cohort of 14 SARS-CoV-2-naïve healthy individuals (average age of 58.8 yrs.) that had received the primary dose followed by a booster after five weeks (Fig. 1b; Supplementary Table 1). We investigated circulating cytokine and antibody levels as well as immune transcriptomes from peripheral blood mononuclear cells (PBMCs) using RNA-seq (Fig. 1c).

**Figure 1.**
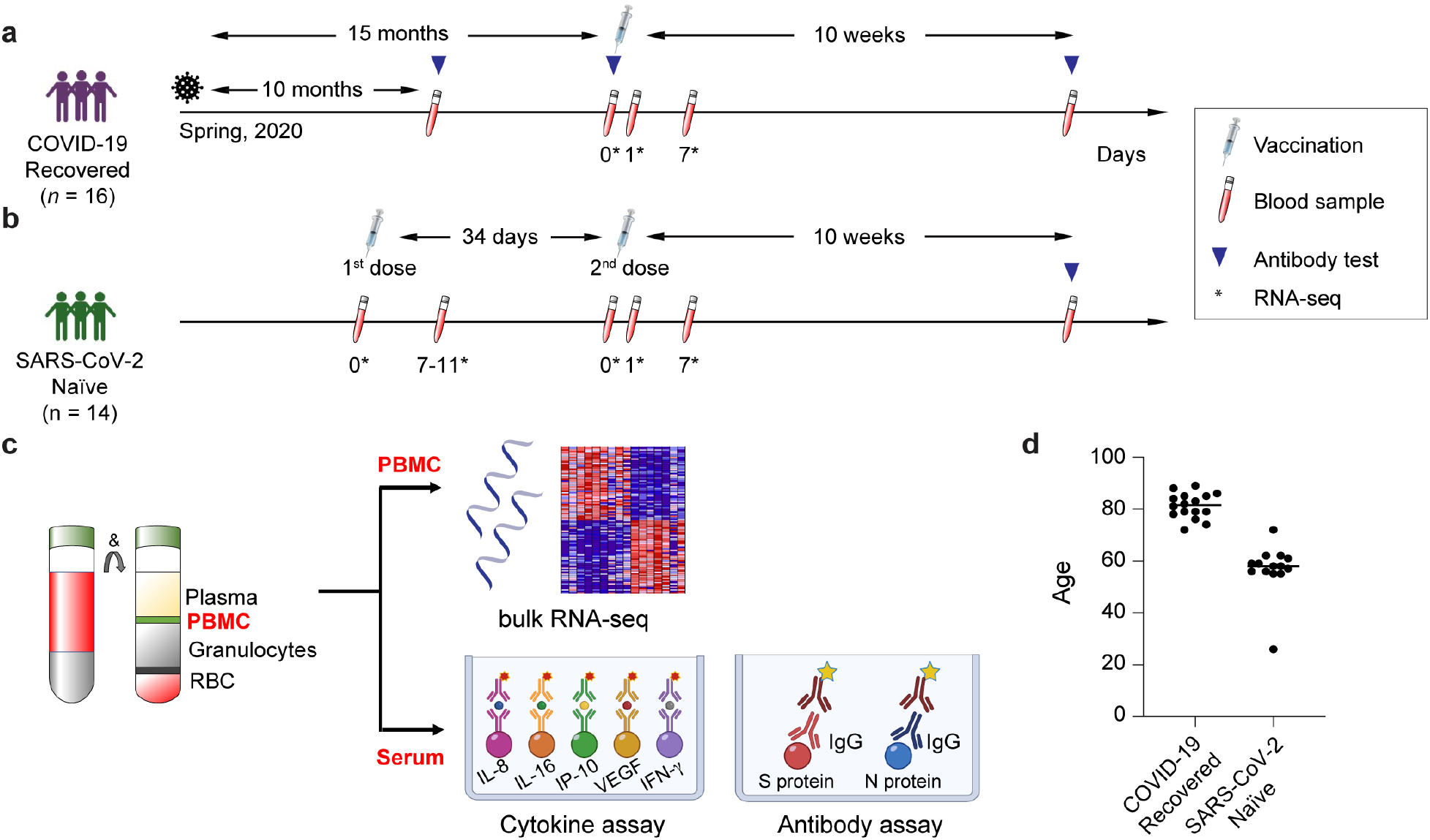
Experimental design. **a-c**. Schematic presentation of the experimental workflow. (a) elderly population recovered from COVID-19; (b) SARS-CoV-2 antigen-naïve population; the syringes indicate vaccinations; the test tubes indicate the blood drawing and the triangles the antibody testing. The asterisks indicate the days at which samples for RNA-seq was collected. (c) Peripheral blood mononuclear cells (PBMCs) were purified from blood drawn prior to and after the vaccination and RNA-seq was performed at a depth of 240 million reads per sample. **d**. Age distribution of participants in each cohort.

### Symptoms and systemic cytokine levels after BNT162b vaccination

Four out of the 16 elderly COVID-19 recovered individuals (Fig. 1d) reported post-vaccine symptoms, fatigue and joint pain, after the primary (booster) dose (Supplementary Table 1). No symptoms were reported by the healthy SARS-CoV-2-naïve individuals after their second (booster) vaccination (Supplementary Table 1). Next, we measured serum levels of a panel of cytokines in the COVID-19 recovered individuals prior to and after the primary booster vaccination, and prior to and after the second (booster) vaccination in the SARS-CoV-2 antigen-naïve group (Fig. 2; Supplementary Table 2). Out of the 10 cytokines measured, an increase of circulating IFNγ and CXCL10 was observed in both groups within one day following vaccination (Fig. 2a-b). IFNγ levels in both groups returned to baseline levels at day 7 (Fig. 2c-d). By day 7, CXCL10 levels had returned to baseline in the naïve group but remained elevated in the COVID-19 recovered group (Fig. 2c-d) suggesting an extended immune response. Levels of the other cytokines tested (IL-2, IL-4, IL-6, IL-8, IL-10, IL-15, IL-16, IL-1β, TNF-α and VEGF) were unchanged in the COVID-19 recovered group (Supplementary Fig. 1). In contrast, in the naïve group, IL-16 levels declined post vaccination and IL-8 levels increased at day 7 post vaccination (Supplementary Fig. 1b).

**Figure 2.**
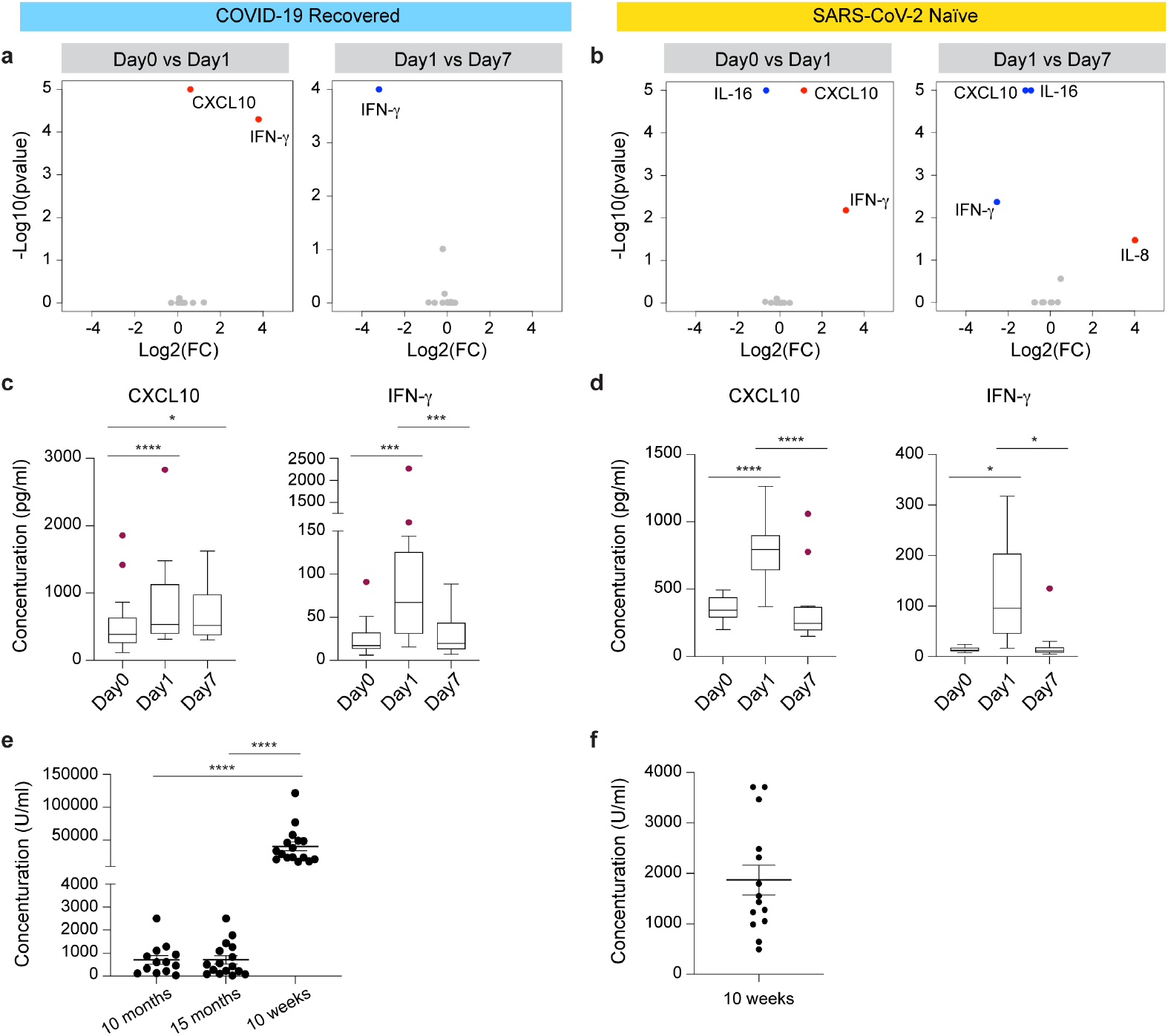
Serum cytokine, chemokine and antibody levels. **a-b**. Volcano plots depict differentially expressed cytokines and chemokine between days 0, 1 and 7 at the 1^st^ vaccination of the COVID-19 recovered cohort (a) and the 2^nd^ vaccination of the SARS-CoV-2 naïve cohort (b). Red dots indicate significant upregulation; blue dots indicate significant downregulation (p-value < 0.05). See also Supplementary Figure 1 and Supplementary Tables 2. **c-d**. Cytokine and chemokine concentrations in serum prior to and post vaccination (pg/ml) of CXCL10 and IFN-γ. Boxes show median, 25th and 75th percentiles and whiskers show the range. p-value are from 2-way ANOVA with Tukey’s multiple comparisons test and Two-stage linear step-up procedure of Benjamini, Krieger and Yekutieli method and listed in Supplementary Table 2. \**p* < 0.05, \*\*\**p* < 0.0001, \*\*\*\**p* < 0.00001. **e-f**. IgG anti-Spike-RBD levels in the COVID-19 recovered (e) and SARS-CoV-2 antigen-naïve group (f). p-value are from 1-way ANOVA with Tukey’s multiple comparisons test. \*\*\*\**p* < 0.00001. Vaccination of the COVID-19 recovered cohort occurred immediately after collecting blood for antibody measurements (15 months).

### Antibody responses to BNT162b vaccination

We first measured circulating antibody responses in serum samples by enzyme-linked immunosorbent assay (ELISA). All elderly COVID-19 recovered individuals had levels of anti-spike IgG ranging from 34 U/ml to >2,500 U/ml (average 706 U/ml) at 10 months after developing COVID-19 (Fig. 2e; Supplementary Table 3). These antibody levels remained largely unchanged five months later (average 710 U/ml), at the day of the primary (booster) vaccination. Anti-spike antibodies were measured again 10 weeks after the vaccination and levels in the 16 individuals ranged from 17,000 to 121,000 U/ml (average 40,599 U/ml) (Fig. 2e; Supplementary Table 3). In contrast antibody levels in the 14 SARS-CoV-2-naïve individuals after the second dose of BNT162b ranged from 500 to 3,700 U/ml (average 2187 U/ml) (Fig. 2f; Supplementary Table 3). This demonstrates an approximately 20-fold elevated response in the elderly, previously infected cohort.

### Transcriptome response of COVID-19 recovered and SARS-CoV-2 antigen-naïve individuals

The exceptional response of the COVID-19 recovered individuals to the booster vaccination promoted us to investigate the molecular innate immune response. We directly compared immune transcriptomes induced by BNT162b vaccination in the COVID-19 recovered cohort and the SARS-CoV-2 antigen-naïve population (Supplementary Tables 4-13). We performed bulk mRNA-seq of PBMCs isolated from the COVID-19 recovered individuals prior to the primary (booster) vaccination and after one and seven days (Fig. 1a). PBMCs from the naïve population were isolated prior to the secondary (booster) vaccination and after one and seven days (Fig. 1b). RNA-seq was conducted on 115 samples with an average of 240 million reads per sample (Supplementary Table 1).

PCA plot analysis indicated that the greatest transcriptome differences in the COVID-19 recovered individuals were found at day 7 post vaccination (Fig. 3a). A total of 161 genes were induced at least two-fold within one day after the booster vaccination (Fig. 3b; Supplementary Table 4), 894 genes were induced by day 7 (Supplementary Table 5) and 652 genes were activated between day 1 and day 7 (Supplementary Table 6). Gene-set enrichment analysis (GSEA) demonstrated that the genes activated within one day after the vaccination were enriched in immune-response, interferon and JAK-STAT pathways (Supplementary Fig. 2). The SARS-CoV-2 naïve group exhibited a distinctly different transcriptome response (Fig. 3c and d). While expression of 173 genes was induced within one day post vaccination (Fig. 3d; Supplementary Table 7), only 32 genes were elevated at day 7 as compared to day 0 (Fig. 3d; Supplementary Table 8). A total of 77 genes were activated between days 1 and 7 (Supplementary Table 9). As expected, the genes activated at day 1 were part of interferon and cytokine pathways (Supplementary Fig. 2d). We also investigated the immune transcriptomes prior to and after the primary vaccination of the SARS-CoV-2 antigen-naïve group and very few induced genes related to chemokine and cytokine signaling were identified (Supplementary Table 10; Supplementary Fig. 3).

**Figure 3.**
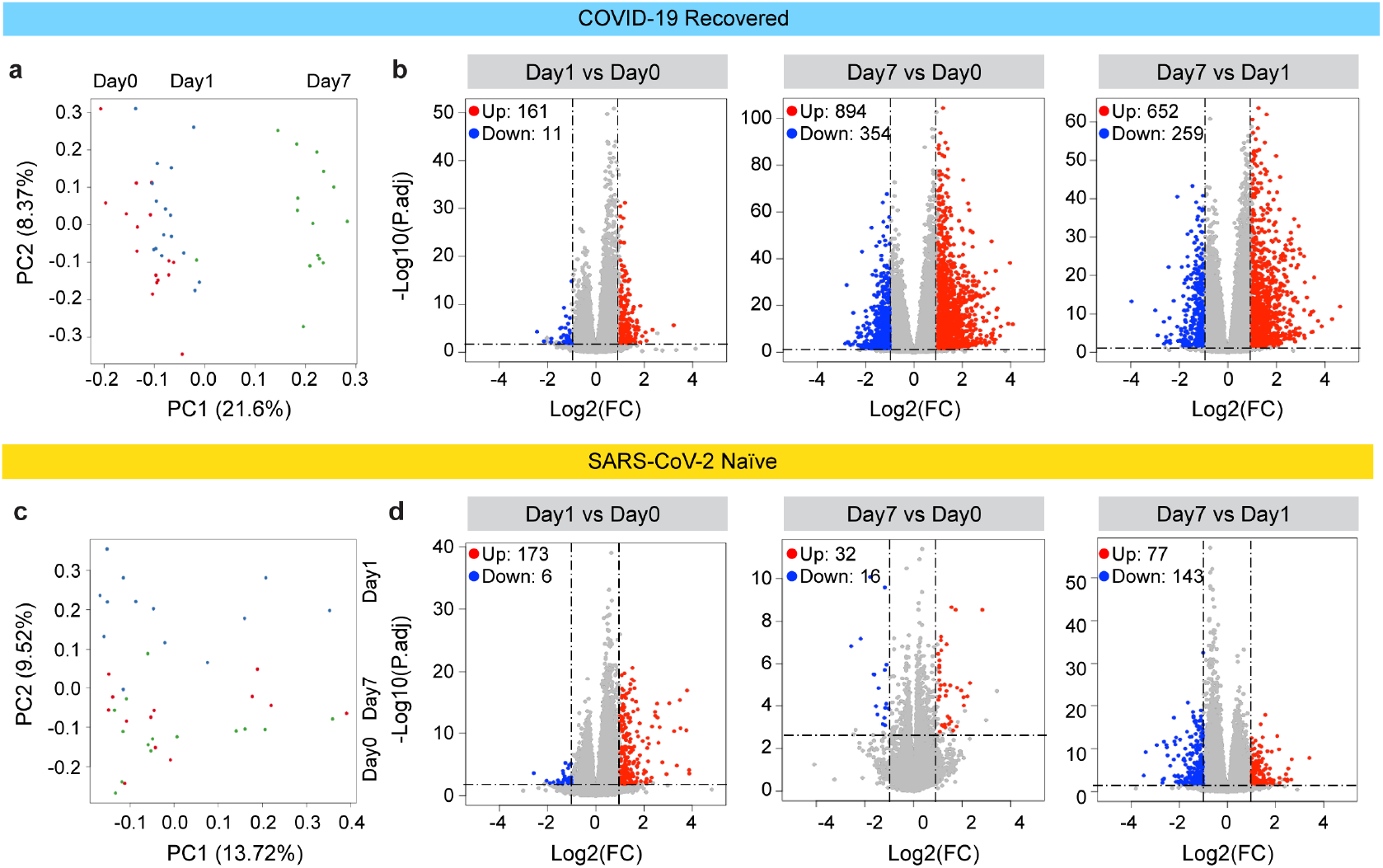
Characteristics of immune transcriptome for vaccination. Principal-component analysis (PCA) of transcriptomes generated prior to (day 0) and after the vaccination (days 1 and 7) from COVID-19 recovered individuals. The variation in the global gene expression profiles across the three time points is shown. Principal components 1 (PC1) and 2 (PC2), which represent the greatest variation in gene expression, are shown. **b**. Volcano plots of DEGs comparing Day1 versus Day0, Day7 vs Day0 and Day7 vs Day1 in the COVID-19 recovered cohort. DEGs (adjusted p-value, P.adj < 0.05) with a log2 fold change (FC) of more than 1 or less than -1 are indicated in red and blue, respectively. Non-significant DEGs are indicated in gray. The numbers of upregulated and downregulated genes are listed in Supplementary Tables 4-6. **c**. PCA of transcriptomes from the SARS-CoV-2 antigen-naïve cohort prior to (day0) and day1 and day7 after the second (booster) vaccination. **d**. Volcano plot of DEGs comparing Day1 versus Day0, Day7 vs Day0 and Day7 vs Day1 in SARS-CoV-2 naïve cohort. The numbers of upregulated and downregulated genes are listed in Supplementary Tables 7-9. Volcano plots depict differentially expressed cytokines and chemokine at day 0, 1 and 7 of 1^st^ vaccination of COVID-19 recovered cohort (a) and 2^nd^ vaccination of SARS-CoV-2 naïve cohort (b). Red dots indicate significant upregulation; blue dots indicate significant downregulation (p-value < 0.05). See also Supplementary Figure 1 and Supplementary Tables 2.

### Differential transcriptome activation between COVID-19 recovered and SARS-CoV-2 naïve individuals

To further understand the stark differences to the booster vaccination between the two groups, we dug deeper and analyzed the longitudinal expression of the genes activated at day 1 post vaccination. Out of the 161 genes activated in the COVID-19 recovered population, expression of 108 genes was still significantly activated at day 7 (Fig. 4a; Supplementary Table 11). Most of these genes are part of interferon and virus-response pathways (Supplementary Table 11). In contrast, out of the 173 genes induced in the naïve population at day 1, only five genes (IFI44, IFI44L, RSAD2, IFIT1 and GBP1P1) were expressed at elevated levels at day 7 (Fig. 4a; Supplementary Table 12). These findings indicate a prolonged vaccine-induced transcriptomic response in COVID-19 recovered individuals.

**Figure 4.**
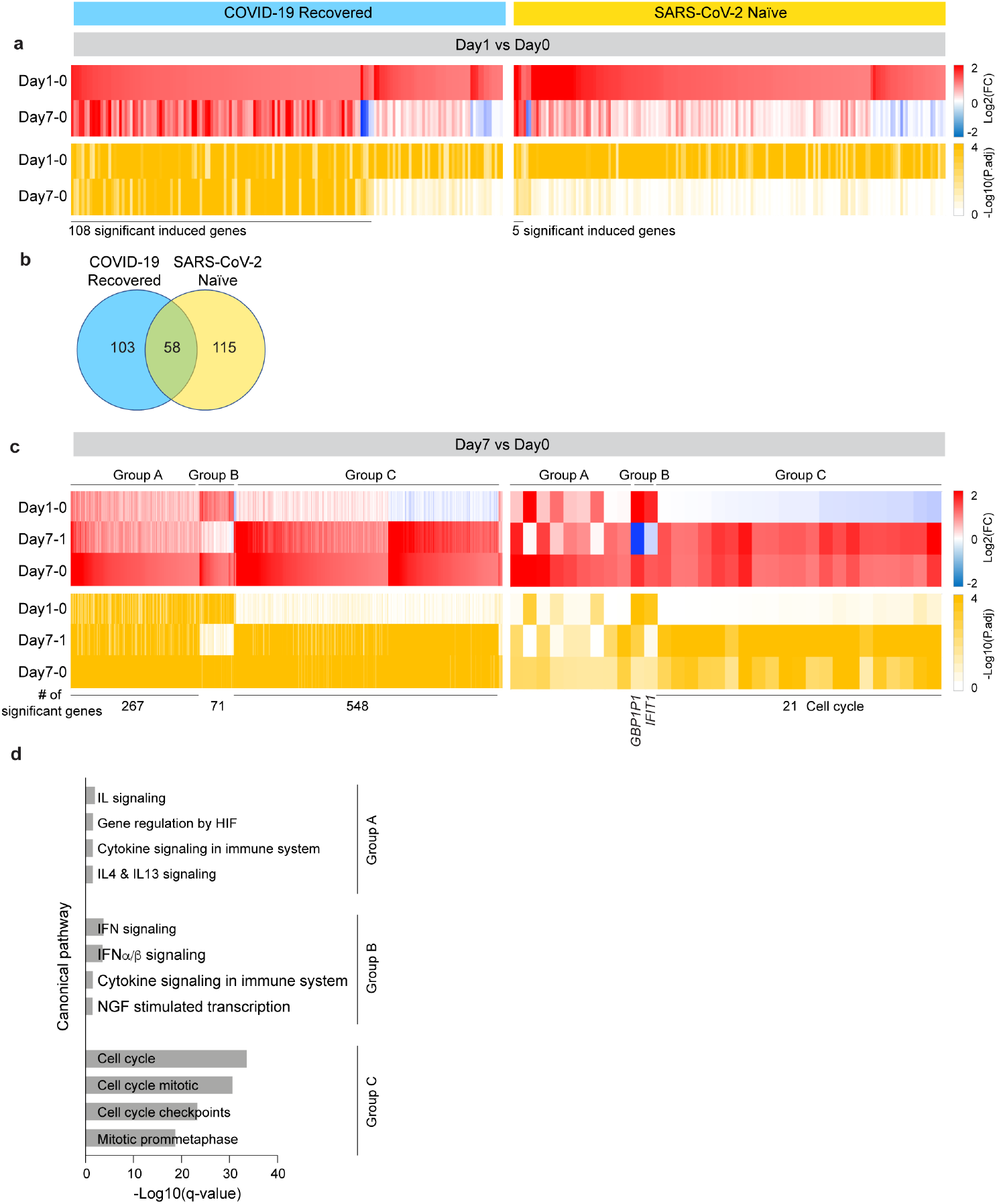
Comparison of immune transcriptomes between COVID-19 recovered and SARS-CoV-2 naïve cohorts after receiving the BNT162b booster. **a**. Heatmaps showing log2 FC (top, red) and corresponding P.adj (bottom, ocher) of significantly upregulated genes between Day0 and Day1 in COVID-19 recovered (left) and SARS-CoV-2 naïve (right) cohorts. The genes specifically activated between day0 and day1 and still induced by day7 are listed in Supplementary Table 11-12. **b**. Venn diagram displays the number of significantly induced genes between Day0 and Day1 in both cohorts. The genes in each part are listed in Supplementary Table 13 and their GSEA analysis is in Supplementary Figure 4. **c**. Heatmaps showing log2 FC (top) and corresponding P.adj (bottom) of genes significantly activated at day7 in COVID-19 recovered (left) and SARS-CoV-2 naïve (right) cohorts. The genes induced between day0 and day7 fall into different categories based on their activation pattern (Supplementary Table 11-12). Group A, genes whose activation progresses between day0 and day7; Group B, genes highly activated preferentially between day0 and day1; Group C, genes highly activated between day1 and day7. **d**. Genes categorized in c were significantly enriched in Hallmark gene sets. X-axis denotes statistical significance as measured by minus logarithm of FDR q-values.

A direct comparison of genes induced in both groups at day 1 post vaccination identified 58 immune relevant genes shared between the COVID-19 recovered and SARS-CoV-2 naïve population (Fig. 4b; Supplementary Table 13; Supplementary Fig. 4). The 115 genes preferentially activated in the naïve population are part of interferon signaling and the 103 genes differentially expressed in the COVID-19 recovered population are enriched for transcriptional events (Supplementary Table 13; Supplementary Fig. 4).

Unlike in the naïve population, extensive persistent transcriptome changes were observed in the COVID-19 recovered population and additional gene classes were activated between days 1 and 7 post vaccination (Fig. 4c-d; Supplementary Table 11; Supplementary Fig. 5). While a total of 548 genes were induced at least two-fold in the COVID-19 recovered cohort, only 21 genes were induced in the SARS-CoV-2 antigen-naïve population between day 1 and 7 post vaccination. Canonical pathways (CP) analysis identified an enrichment in cell cycle pathway genes in the COVID-19 recovered population (Fig. 4d).

### Shared transcriptomes induced by different vaccines

Transcriptome (RNA-seq) studies have been reported for the hepatitis B vaccine^13^, influenza vaccines^14,15^ and the BNT162b vaccine in a younger cohort (avg. 39 yrs.)^16^. To understand transcriptome responses to different vaccines, we have compared genes reported to be activated in the published studies within 1-2 days post vaccination (Supplementary Fig. 6). The immediate (day0) response to the BNT162b booster in our Naïve cohort was similar to that induced by the inactivated influenza vaccine and mRNA BNT162b2 vaccine. In contrast only five genes identified in the Hepatitis B vaccine study (Supplementary Fig. 6). However, since the sequencing depth in these studies is significantly lower (∼38 million reads per sample^16^ as compared to 240 million in our study) a definitive direct comparison is challenging.

## Discussion

In this real-world study we provide evidence that a single dose of the mRNA vaccine BNT162b elicits a strong immune response in an elderly population vaccinated 15 months after recovering from COVID-19. Aging is associated with a decline of the immune system, commonly referred to as immunosenescence, and increased chronic low-grade systemic inflammation, also referred to as inflammaging^17^, both coupled with a poor vaccine response^8^. However, our data demonstrate that the immune response to BNT162b in an elderly population (avg. 81 yrs.) previously infected with SARS-CoV-2 exceeds that of a younger SARS-CoV-2 naïve cohort (avg. 59 yrs.) receiving the two-dose regimen.

The optimal window for providing the booster vaccine to individuals recovered from a previous SARS-CoV-2 infection has not been defined and might be age-dependent. Recent studies have investigated the immune response in younger populations recovered from COVID-19^10,11,18-20^. In general, the immune responses, including spike-specific IgG antibody levels, in individuals younger than 50 years having received booster doses within one to six months after the original SARS-CoV-2 infection were similar to those seen after two doses of vaccine in individuals of similar age without prior infection^10,11,18,21^. A large scale clinical study provided evidence that natural immune protection that develops after a SARS-CoV-2 infection followed by a single booster vaccination provides considerably more protection against the SARS-CoV-2 Delta variant than two doses of the Pfizer-BioNTech vaccine in SARS-CoV-2 naïve individuals^20^. It is still not clear if one or two boosters are indicated for previously infected individuals and different countries are advocating different approaches with some dictating one booster and others two. Although one booster vaccination appears to result in a solid immune response in COVID-19 recovered individuals, studies with two boosters are emerging^21^. Third booster vaccinations are now available for SARS-CoV-2 naïve individuals (https://www.cdc.gov/coronavirus/2019-ncov/vaccines/booster-shot.html) and its effectiveness in reducing transmission and severe disease was demonstrated in individuals 60 years and older who had been fully vaccinated the two-dose regimen at least five months previously^17^. Longitudinal antibody measurements in our elderly cohort will provide a better understanding on the need and timing of a second booster dose in this vulnerable population.

Our study shows a strong, prolonged immune response in the COVID-19 recovered elderly that have received one booster vaccination, greatly exceeding that seen after the booster vaccination in SARS-CoV-2 naïve individuals. This is seen both in generation of antibody and in transcriptomes. Exploring the genomic immune responses after vaccination through RNA-seq approaches can identify transcriptional signatures associated with effective antibody production but published data is limited to younger age individuals (avg 39 yr.)^13,16^ Sequencing depth is a consideration for interpretation of such studies. Both ours and the study conducted in younger individuals^16^ focused on the response to the Pfizer-BioNTech vaccine. The younger age study^16^, using an average sequencing depth of approximately 38 million reads per sample, was sufficient for identifying antiviral and interferon signatures, but could not identify significant differences in gene signatures. In contrast, our study, utilizing a minimum of 240 million reads for each sample, was able to identify the specific gene signatures that underlie viral and interferon response. This enabled us to characterize a temporally extended and expanded genetic responses induced by a single BNT162b vaccine in the elderly with prior COVID 19 infection and extend knowledge on the response of SARS-CoV-2 naïve individuals after the second vaccination.

Results from this real-world study are encouraging for vaccine efficacy in the elderly previously infected with SARS-CoV-2. Antibody levels in this group greatly exceeded those in a younger cohort receiving the two-dose regimen. While the optimal window between previous infection and booster shot is not known, our study demonstrates that a 15-months gap did not negatively interfere with the immune response but resulted in a robust production of antibodies. This has practical implications for health care professionals making decisions on the need to booster vaccinations.

### Limitations of this study

This elderly cohort previously infected with SARS-CoV-2 was from a narrowly defined geographically area and included only one gender (females). The infection occurred in the spring of 2020 and the viral strain had not been sequenced. The SARS-CoV-2 antigen-naïve population was from a narrowly defined geographically area. The study was confined to the BNT162b mRNA vaccine.

## Methods

### Study population, study design and recruitment

Sixteen COVID-19 recovered volunteers who were infected with SARS-CoV-2 and developed COVID-19 in the spring of 2020 and 14 SARS-CoV-2 naïve healthy volunteers and (Supplementary Table 1) were recruited for the study under informed consent. Recruitment and blood sample collection took place between January and August 2021. This study was approved (EK Nr: 1064/2021) by the Institutional Review Board (IRB) of the Office of Research Oversight/Regulatory Affairs, Medical University of Innsbruck, Austria, which is responsible for all human research studies conducted in the State of Tyrol (Austria). The investigators do not need to have an affiliation with the University of Innsbruck. A waiver of informed consent was obtained from the Institutional Review Board (IRB) of the Office of Research Oversight/Regulatory Affairs, Medical University of Innsbruck (https://www.i-med.ac.at/ethikkommission/). Written informed consent was obtained from all subjects. This study was determined to impose minimal risk on participants. All methods were carried out in accordance with relevant guidelines and regulations. All research has been have been performed in accordance with the <u>Declaration of Helsinki</u> (https://www.wma.net/policies-post/wma-declaration-of-helsinki-ethical-principles-for-medical-research-involving-human-subjects/). In addition, we followed the <u>‘Sex and Gender Equity in Research – SAGER – guidelines’</u> and included sex and gender considerations where relevant.

### Quantification of immunoproteins

Serum samples from all participants were collected from their blood. After thawing, serum samples were centrifuged for 3 minutes at 2000 *g* to remove particulates prior to sample preparation and analysis. The electrochemiluminescence V-PLEX assay (Meso Scale Discovery, MD) was used to measure proinflammatory proteins (IFN-γ, IL-1β, IL-2, IL-4, IL-6, IL-8, IL-10 and TNF-a), cytokines (IL-15, IL16 and VEGF) and chemokine (CXCL10). Serum samples were diluted 2-fold and measured in duplicate. The cytokines concentration was determined with the electrochemiluminescent labels whilst the plate is inserted into the MSD instrument (MESO QUICKPLEX SQ 120). All samples were assayed in duplicate. High and low controls were used to assess variance between plates. The inter-assay coefficient of variations was <10%. The results were analyzed using MSD DISCOVERY WORKBENCH analysis software.

### Anti-S binding ELISA

Anti-SARS-CoV-2 S-Protein antibodies were measured using the Roche Elecsys Anti-SARS-CoV-2 S immunoassay on a Cobas e411 analyzer (Roche Diagnostics, Basel, Switzerland) as described somewhere else^18^. In short, serum samples were incubated with biotinylated and adenylated recombinant SARS-CoV-2 RBD antigen. In presence of corresponding anti-SARS-CoV-2 antigens immune complexes were formed, which were bound to the solid phase after addition of streptavidin-coated microparticles. Microparticle-bound antibodies were detected by electrochemiluminescence in the Cobas e411 analyzer. Results <0.8 U/mL were diagnosed as non-reactive, while results >0.8 U/mL were diagnosed as reactive.

### Extraction of the buffy coat and purification of RNA

Whole blood was collected, and total RNA was extracted from the buffy coat and purified using the Maxwell RSC simply RNA Blood Kit (Promega) according to the manufacturer’s instructions. The concentration and quality of RNA were assessed by an Agilent Bioanalyzer 2100 (Agilent Technologies, CA).

### mRNA sequencing (mRNA-seq) and data analysis

The Poly-A containing mRNA was purified by poly-T oligo hybridization from 1 μg of total RNA and cDNA was synthesized using SuperScript III (Invitrogen, MA). Libraries for sequencing were prepared according to the manufacturer’s instructions with TruSeq Stranded mRNA Library Prep Kit (Illumina, CA, RS-20020595) and paired-end sequencing was done with a NovaSeq 6000 instrument (Illumina) yielding 200-350 million reads per sample.

The raw data were subjected to QC analyses using the FastQC tool (version 0.11.9) (https://www.bioinformatics.babraham.ac.uk/projects/fastqc/). mRNA-seq read quality control was done using Trimmomatic ^19^ (version 0.36) and STAR RNA-seq ^20^ (version STAR 2.5.4a) using 150 bp paired-end mode was used to align the reads (hg19). HTSeq ^21^ (version 0.9.1) was to retrieve the raw counts and subsequently, Bioconductor package DESeq2 ^22^ in R (https://www.R-project.org/) was used to normalize the counts across samples and perform differential expression gene analysis. Additionally, the RUVSeq ^23^ package was applied to remove confounding factors. The data were pre-filtered keeping only genes with at least ten reads in total. The visualization was done using dplyr (https://CRAN.R-project.org/package=dplyr) and ggplot2 ^24^. The genes with log2 fold change >1 or <-1 and adjusted p-value (pAdj) <0.05 corrected for multiple testing using the Benjamini-Hochberg method were considered significant and then conducted gene enrichment analysis (GSEA, https://www.gsea-msigdb.org/gsea/msigdb).

### Statistical analysis

Differential expression gene (DEG) identification used Bioconductor package DESeq2 in R. P-values were calculated using a paired, two-side Wilcoxon test and adjusted p-value (pAdj) corrected using the Benjamini–Hochberg method. Genes with log2 fold change >1 or <-1, pAdj <0.05 and without 0 value from all sample were considered significant. For significance of each GSEA category, significantly regulated gene sets were evaluated with the Kolmogorov-Smirnov statistic. P-values of cytokines were calculated using two-stage linear step-up procedure of Benjamini, Krieger and Yekutieli on GraphPad Prism software (version 9.0.0). A value of \**P* < 0.05, \*\**P* < 0.001, \*\*\**P* < 0.0001, \*\*\*\**P* < 0.00001 was considered statistically significant.

## Data Availability

The RNA-seq data from this study will be uploaded in GEO before publishing the manuscript.

## Ethics approval

This study was approved (EK Nr: 1064/2021) by the Institutional Review Board (IRB) of the Office of Research Oversight/Regulatory Affairs, Medical University of Innsbruck, Austria, which is responsible for all human research studies conducted in the State of Tyrol (Austria) regardless of whether or not the investigators have an affiliation with the University of Innsbruck.

## Data availability

The RNA-seq data from this study will be uploaded in GEO before publishing the manuscript.

## Acknowledgments

Our gratitude goes to the participants who contributed to this study to advance our understanding of COVID-19 vaccination. This work was supported by the Intramural Research Programs (IRPs) of National Institute of Diabetes and Digestive and Kidney Diseases (NIDDK) and utilized the computational resources of the NIH HPC Biowulf cluster (http://hpc.nih.gov). RNA-sequencing was conducted in the NIH Intramural Sequencing Center, NISC (https://www.nisc.nih.gov/contact.htm).

## Author contribution

HKL: generated and analyzed data, wrote manuscript; LK: recruited patients, collected material, analyzed data; LK Sr.: recruited patients, collected material, analyzed data; SK: recruited patients, collected material, analyzed data; BP: recruited patients, collected material, analyzed data; MW: collected and prepared material; PAF: analyzed data, wrote manuscript; LH: analyzed data, wrote manuscript. All authors read and approved the manuscript.

